# Genetic contributions to BMI fluctuation and its associations with BMI and its trajectories over adolescence and early adulthood: a 25-year follow-up longitudinal study of Finnish twins

**DOI:** 10.1101/2025.08.11.25333125

**Authors:** Alvaro Obeso, Aline Jelenkovic, Jose Angel Peña Garcia, Gabin Drouard, Sari Aaltonen, Jaakko Kaprio, Karri Silventoinen

## Abstract

**Objective:** We examined how BMI, BMI trajectories, and BMI fluctuation around these trajectories in adolescence were correlated with BMI trajectories and BMI fluctuation in early adulthood, as well as the genetic basis of these associations.

**Methods:** BMI data from Finnish twins (N=1379, 48% males) were collected at ages 11.5, 14, 17.5, 24, and 37 years. BMI trajectories in adolescence (11.5–17.5 years) and early adulthood (17.5–37 years) were estimated using linear mixed-effect models. BMI fluctuation was calculated as the average squared differences between observed and expected BMI around these trajectories. Genetic twin models and a polygenic risk score for BMI (PRS_BMI_) were used to assess genetic contributions to BMI fluctuation and its associations with BMI and BMI trajectories.

**Results:** Adolescent BMI fluctuation was positively correlated with early adulthood BMI trajectories in females, while in males, adolescent BMI trajectories were positively associated with BMI fluctuation in early adulthood. Genetic factors affected BMI fluctuation in both adolescence and early adulthood when estimated using twin modelling and PRS_BMI_. Adolescent BMI was positively associated with early adulthood fluctuation in both sexes, with genetic factors playing a role (genetic correlations (r_A_) = 0.08 – 0.29).

**Conclusion:** Genetic factors play a significant role in BMI fluctuations in adolescence and early adulthood, with some overlap with the genetics of BMI.

## Introduction

Obesity is considered a major public health challenge across all age groups. A particular concern is childhood obesity, as it is a major risk factor for developing obesity in adulthood (1). While obesity is defined as the excessive accumulation of adipose tissue due to an imbalance between energy intake and expenditure, it is a multifactorial condition influenced by genetics, environment, behavior, and their mutual interactions. The primary indicator of obesity in epidemiological studies is body mass index (BMI), and it is also commonly used in clinical settings to identify individuals who may require further assessment for fat accumulation. The heritability of BMI ranges from 40% to 70% in twin studies (2, 3) and 20% to 50% in family and adoption studies (4, 5).

Additionally, cross-sectional genome-wide association (GWA) studies have identified over 1,000 independent loci associated with adult BMI (6, 7, 8) and several loci associated with BMI in childhood (9, 10). Furthermore, previous studies have shown that genetic factors play an important role in determining BMI trajectories in adulthood (11, 12, 13, 14). However, there are no previous studies about the underpinning genetics of BMI trajectories in adolescence and BMI fluctuations both in adolescence and adulthood.

The role of BMI in adolescence in the development of adult obesity (15) and BMI trajectories in adulthood has been demonstrated in previous studies (14). However, despite extensive research on BMI, only a few previous studies have examined the fluctuation of BMI over time (16) and its associations with different health-related conditions, such as metabolic syndrome (17) and cardiometabolic disease risk (18). BMI fluctuation has important public health and clinical implications since frequent weight cycling can predispose individuals to future weight gain (19, 20). Furthermore, little is still known about the role of genetic and environmental factors in BMI fluctuation and its associations with BMI and its trajectory. Moreover, the association between BMI fluctuation and genetic predisposition to high BMI, as well as the heritability of BMI fluctuation, remains underexplored.

We aimed to analyse the genetic underpinnings of fluctuations in BMI, defined as the temporal variability of BMI around an individual’s fitted BMI trajectory, and how it relates to future BMI and its trajectory over time. Specifically, we analysed the association between (i) BMI in adolescence and BMI fluctuation in early adulthood (ii) BMI trajectories in adolescence and BMI fluctuation in early adulthood (iii) BMI fluctuation in adolescence and BMI trajectories in early adulthood, and (iv) BMI fluctuation in adolescence and BMI fluctuation in early adulthood. Additionally, we investigated whether genetic predisposition to high BMI is associated with BMI fluctuation and BMI trajectories in adolescence and early adulthood. For this, we used two analytical designs, genetic twin modelling and polygenic risk score of BMI (PRS_BMI_), each with different background assumptions, and applied them to longitudinal data. This approach allowed us to unravel the complex interplay between genetics and the environment, as well as account for confounding factors.

## Methods

### Cohort

The data was obtained from the longitudinal FinnTwin12 study, which included five waves of data collection. The initial mailed questionnaire survey (wave 1) was sent to all twins born in Finland between 1983 and 1987 when they were 11 – 12 years old, resulting in a total of 5,362 participants (response rate 87%). Four follow-up surveys (waves 2 – 5) were sent at the average ages of 14 (response rate 88%), 17.5 (response rate 92%), 24 (response rate 66%) and 37 (response rate 43%) (21, 22, 23). Participants reported their current weight and height in each survey, which was used to calculate BMI (kg/m^2^). The correlation between self-reported and measured BMI was 0.97 in a subsample of these twins at the age of 24, indicating good reliability of self-reported BMI (24). Zygosity was assessed using DNA, or if not available, based on questions about physical similarity in the baseline questionnaire. In a validation study of 395 same-sex twin pairs in this same cohort, 97% of questionnaire assignments of zygosity were confirmed by a DNA test, showing high reliability (25). The Ethics Committee of the Helsinki University Central Hospital District (HUS) approved the most recent data collection (wave 5) (HUS/2226/2021, dated 22 September 2021), as well as the use of previously collected data. The participants or their parents/legal guardians provided informed consent when participating in the surveys.

Participants with valid BMI measures in all five waves (N=1379) were selected. From these participants, we selected complete twin pairs (N=336 pairs) for genetic twin modelling (Figure 1). Although BMI was not a criterion for selecting twins, we found that the BMI of those selected in the current study was slightly lower, but not statistically significantly, compared to those not included (0.16 – 0.58 kg/m^2^ after adjusting for age and sex in each survey), suggesting that there may be some self-selection related to BMI.

**Figure 1:**
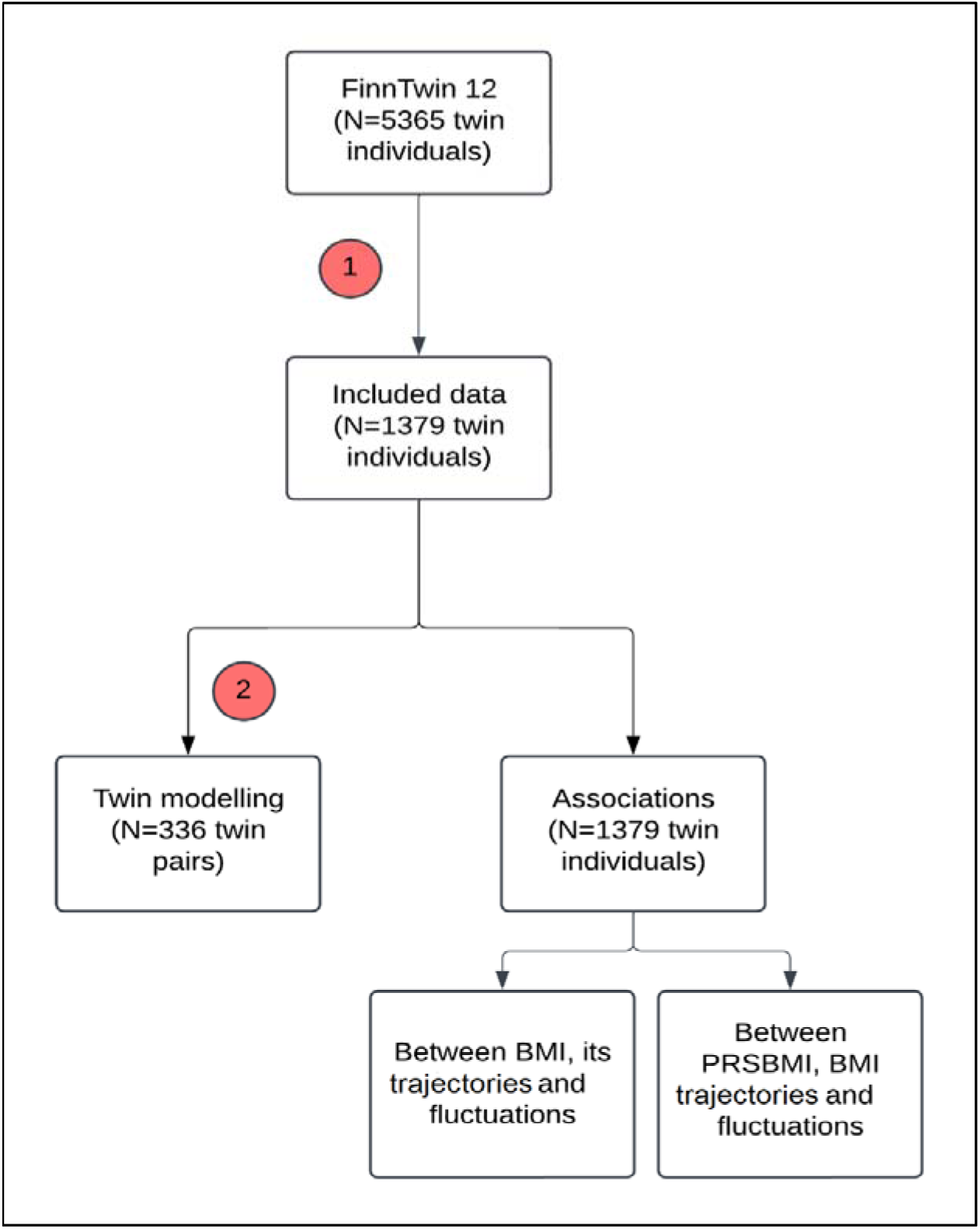
Participant flowchart and general study plan diagram. The study was divided into two main parts based on the analyses after a data preprocessing phase. Preprocessing consisted of obtaining sex-specific BMI trajectories (slope and intercept) and fluctuation from those participants with height and weight measures for the five waves. One part of the study was phenotypic associations between BMI variables themselves and with the polygenic risk score of BMI. The other main part of the study was the classical twin genetic modelling which was based on BMI trajectories (slope and intercept) and fluctuation. Two exclusion criteria were performed at different times: they correspond to (1) data preprocessing and (2) selection of complete same-sex twin pairs. **Abbreviations:** MZ, monozygotic twins; DZ, dizygotic twins; N, number of participants

### BMI trajectory calculation

The BMI trajectories in adolescence were calculated using BMI measures from waves 1 – 3 (between 11.5 and 17.5 years), while for early adulthood BMI trajectories, measures from waves 3 – 5 (from age 17.5 to 37) were used. In both cases, the trajectories were calculated using linear mixed-effect (LME) models that included both fixed and random effects, as explained in detail elsewhere (14). Random effects were applied to individual identifiers to estimate individual baseline BMI (i.e., intercept) and BMI trajectory (i.e., slope). R software (version 4.2.3) packages lme4 (version 1.1-34), lmerTest (version 3.1-3), dplyr (version 1.1.4), modelsummary (version 1.4.3) and optimx (version 2023-10.21) were used.

### BMI fluctuation calculation

The participant-specific BMI trajectories and baseline BMI were used to estimate BMI fluctuation. Initially, the BMI trajectory value was determined by using the formula *y = xn + m*, where *n* represents the BMI trajectory measured in kg/m^2^ per year, *m* is the baseline BMI measured in kg/m^2^ (i.e., wave 1 for adolescence and wave 3 for early adulthood), and *x* is the difference between the age of interest and the baseline age (i.e., wave 1 for adolescence and wave 3 for early adulthood). Next, the squared differences between observed and expected BMI values were calculated at each age. Finally, the total BMI fluctuation in adolescence and early adulthood were calculated for each participant by averaging the squared difference values (using waves 1, 2, and 3 for adolescence and waves 3, 4, and 5 for early adulthood). A sensitivity analysis was conducted to evaluate the accuracy of the method used for the estimation of BMI fluctuation. For that, another method (one that makes no assumptions about the linearity of BMI trajectories) was used for estimation of BMI fluctuation and then it was compared to the previously used method (26). A strong correlation was found between the BMI fluctuations obtained in these different ways (Supplementary Table 1 and related text).

### Polygenic risk score of high BMI (PRS_BMI_)

At wave 4, DNA from a subset of twins selected from the cohort independently of their BMI was obtained from venous blood (for twins who attended the young adult assessment in person) or saliva samples (using Oragene kits mailed along with the instructions of a study protocol that could be completed from home and which was an alternative offered to twins who had not taken part in the studies conducted at the Department of Public Health in 2006-2009 in Helsinki). These individuals (N=1,478) were used in the PRS_BMI_ analyses (22). The technical details of genotyping, quality control and imputation in genotype data processing have been described elsewhere (27). The PRS_BMI_ was derived using GWA summary statistics from Yengo et al. (2018) (8). For the calculation of the PRS_BMI_, a total of 996,919 single nucleotide polymorphisms (SNPs) with a minor allele frequency >5% in European individuals obtained from 692,578 participants were used. To correct for population stratification, the PRS_BMI_ was regressed to the top ten genetic principal components, and the residuals were scaled to a mean of zero and a variance of one (28). We found no difference in PRS_BMI_ distributions between included and excluded participants (p=0.19), suggesting that there was no selection bias for PRS_BMI_.

### Association study

Linear mixed-effects regression models were carried out to calculate the associations between (i) BMI in adolescence (at waves 1, 2 and 3) and BMI fluctuation in early adulthood (ii) BMI trajectories in adolescence and BMI fluctuation in early adulthood (iii) BMI fluctuation in adolescence and BMI trajectories in early adulthood, and (iv) BMI fluctuation in adolescence and BMI fluctuation in early adulthood. (the models used are displayed in a footnote of Table 3). Furthermore, associations between the PRS_BMI_ and BMI fluctuation in adolescence and early adulthood were assessed to determine whether genetic predispositions for high BMI were associated with higher BMI fluctuation in adolescence and early adulthood (the models used for the study of associations of PRS_BMI_ with BMI trajectories and fluctuations in adolescence and early adulthood are displayed in a footnote of Table 4). All analyses were conducted separately for males (n=665) and females (N=714), since mean BMI (from wave 3 onwards) and BMI trajectories differed between males and females. We considered associations 0 – 0.39 as weak, 0.40 – 0.59 as moderate and 0.60 – 1.00 as strong. The analyses were conducted using the R software (version 4.2.3) and the R package lme4 (version 1.1-35.5) and lmerTest (version 3.1-3).

### Genetic twin modelling

The genetic analyses were continued by twin modelling based on the different genetic relatedness of monozygotic (MZ) and dizygotic (DZ) twins: while MZ twins have virtually identical genetic sequences, DZ twins share, on average, 50% of their genes identical-by-descent. The trait variance can be decomposed into four components: (i) additive genetic variation (A), which includes the effects of all the loci that play a relevant role in the trait (correlation of 1 in MZ twins and 0.5 in DZ twins); ii) effects due to dominance (D) where the heterozygote phenotype deviates from the additive model; (iii) shared environmental component (C), including all the environmental factors that make the co-twins similar (correlation of 1 in MZ twins and 1 in DZ twins), and (iv) unique environmental component (E), including all environmental factors making the co-twins different along with the measurement error (correlation of 0 in MZ and DZ twins). For twins reared together, all four components cannot be simultaneously estimated, hence ACE and ADE are chosen as starting models depending on the pattern of MZ and DZ correlations. ACE models are used when the MZ twin pair correlations (rMZ) are at least twice as large as the DZ twin pair correlation (rDZ). If the correlations of MZ twin pairs are less than two times the correlation of DZ twins (rMZ>2rDZ), ADE models are estimated.

Separate univariate models for each trait were used to determine the best fitting model and to calculate the relative contributions of genetic and environmental factors. Intra-class correlations by zygosity were calculated by dividing within-pair variation by between-pair variation using the analysis of variance. Based on these correlations, the additive genetic/shared environment/unique environment (ACE) model was selected as the baseline model (Supplementary table 2). Model comparisons were conducted using - 2 log likelihood (-2LL) and Akaike information criterion. Based on the model comparisons, a full AE model with sex differences was selected for all the variables (Supplementary table 3). However, in females the model suggests the presence of a shared environmental component (C) having rDZ > rMZ. However, this result can be attributed to small sample size.

Finally, the bivariate Cholesky decomposition, which is a model-free method to decompose all variation and covariation in the data into uncorrelated latent factors (26), was used to decompose the covariation between the different measures into genetic and environmental covariances. When these covariances are standardised, we obtain estimates of additive genetic (r_A_) and unique environmental (r_E_) correlations. Genetic twin modelling was performed using the R software (version 4.2.3) and the R package OpenMx (version 2.21.11). The 95% confidence intervals (CI) were assessed using maximum likelihood estimation (27).

## Results

Table 1 displays the descriptive information of the data used in the current study. Mean BMI increased from 17.48 kg/m^2^ in males and 17.39 kg/m^2^ in females in wave 1 to 26.09 kg/m^2^ and 24.93 kg/m^2^, respectively, in wave 5. Males showed a higher mean BMI from age 17.5 onwards, which coincided with the establishment of statistically significant BMI differences between the sexes (p<0.01). Accordingly, statistically significant sex differences were observed for the estimates of BMI trajectories, being also higher in males in adolescence (0.64 kg/m^2^ per year in males and 0.54 kg/m^2^ per year in females, p<0.01) and early adulthood (0.22 kg/m^2^ per year in males and 0.20 kg/m^2^ per year in females, p=0.01).

**Table 1.** Means and standard deviations of body mass index (BMI) at different waves and its trajectories in adolescence (waves 1 to 3) and early adulthood (from wave 3 to 5) by sex.

|  | Males (n=665) |  | Females (n=714) |  | Sex differences (p value) |
| --- | --- | --- | --- | --- | --- |
|  | Mean | SD | Mean | SD |  |
| **Wave 1** |  |  |  |  |  |
| Age (years) | 11.41 | 0.30 | 11.42 | 0.30 | 0.79 |
| BMI (kg/m<sup>2</sup>) | 17.48 | 2.39 | 17.39 | 2.45 | 3.91e-07 |
| **Wave 2** |  |  |  |  |  |
| Age (years) | 14.03 | 0.07 | 14.03 | 0.06 | 0.30 |
| BMI (kg/m<sup>2</sup>) | 19.20 | 2.56 | 19.24 | 2.44 | 0.91 |
| **Wave 3** |  |  |  |  |  |
| Age (years) | 17.60 | 0.20 | 17.59 | 0.20 | 0.17 |
| BMI (kg/m<sup>2</sup>) | 21.42 | 2.47 | 20.81 | 2.41 | <2.2e-16 |
| **Wave 4** |  |  |  |  |  |
| Age (years) | 24.14 | 1.70 | 24.19 | 1.62 | 0.06 |
| BMI (kg/m<sup>2</sup>) | 23.90 | 3.06 | 22.36 | 3.27 | <2.2e-16 |
| **Wave 5** |  |  |  |  |  |
| Age (years) | 37.13 | 1.42 | 37.23 | 1.47 | 0.02 |
| BMI (kg/m<sup>2</sup>) | 26.09 | 3.75 | 24.93 | 4.24 | <2.2e-16 |
| **BMI trajectories<sup>1</sup>** |  |  |  |  |  |
| BMI adolescence trajectories (kg/m<sup>2</sup> per year) | 0.64 | 0.15 | 0.54 | 0.12 | <2.2e-16 |
| BMI early adulthood trajectories (kg/m<sup>2</sup> per year) | 0.22 | 0.08 | 0.20 | 0.12 | 5.34e-12 |

<sup>1</sup>The BMI trajectory estimates were obtained by linear regression models based on BMI at waves 1, 2 and 3 for adolescence, and at waves 3, 4, and 5 for early adulthood. The sex difference was studied using linear mixed-effect models: (1) for ages: Age at n wave = Sex + Zygosity + (1| Family ID)). (2) For BMI: BMI at n wave = Age at n wave + Sex + Zygosity + (1| Family ID). (3) For BMI trajectories in adolescence: BMI trajectories in adolescence = BMI intercept for adolescence + Adolescence age baseline + Zygosity + Sex + (1|Family ID). (4) For BMI trajectories in early adulthood: BMI trajectories in adulthood = BMI intercept for adulthood + Adulthood age baseline + BMI intercept for adolescence + Zygosity + Sex + (1| Family ID).
Abbreviations: SD, standard deviation; BMI, body mass index

Table 2 presents the heritability estimates for the BMI trajectories and BMI fluctuation in adolescence and early adulthood for males and females. The heritability of BMI fluctuation in males was higher in adolescence (a^2^=0.80) than in early adulthood (a^2^=0.34), while in females the heritability was higher in early adulthood (a^2^=0.51) than in adolescence (a^2^=0.34). Regarding BMI trajectories, heritability in males was higher in early adulthood (a^2^=0.70) than in adolescence (a^2^=0.61), while in females the heritability was higher in adolescence (a^2^=0.75) compared to early adulthood (a^2^=0.61). However, not all differences were statistically significant, as indicated by overlapping 95% CIs.

**Table 2.** Relative proportions of body mass index (BMI) variance explained by additive genetic and unique environmental variance components with 95% confidence intervals (CI) of BMI trajectories and fluctuation in adolescence and early adulthood by sex.^1^.

|  | Males |  |  |  |  |  | Females |  |  |  |  |  |
| --- | --- | --- | --- | --- | --- | --- | --- | --- | --- | --- | --- | --- |
|  | Additive genetic factors |  |  | Unique environmental factors |  |  | Additive genetic factors |  |  | Unique environmental factors |  |  |
|  | a<sup>2</sup> | 95% CI |  | e<sup>2</sup> | 95% CI |  | a<sup>2</sup> | 95% CI |  | e<sup>2</sup> | 95% CI |  |
|  |  | LL | UL |  | LL | UL |  | LL | UL |  | LL | UL |
| BMI fluctuation in adolescence | 0.80 | 0.69 | 0.87 | 0.20 | 0.13 | 0.31 | 0.34 | 0.17 | 0.49 | 0.66 | 0.51 | 0.83 |
| BMI fluctuation in early adulthood | 0.35 | 0.11 | 0.53 | 0.65 | 0.47 | 0.89 | 0.51 | 0.38 | 0.62 | 0.49 | 0.38 | 0.62 |
| BMI trajectory in adolescence | 0.62 | 0.44 | 0.74 | 0.38 | 0.26 | 0.56 | 0.75 | 0.65 | 0.83 | 0.25 | 0.17 | 0.35 |
| BMI trajectory in early adulthood | 0.71 | 0.56 | 0.80 | 0.29 | 0.20 | 0.44 | 0.61 | 0.47 | 0.72 | 0.39 | 0.28 | 0.53 |

<sup>1</sup>AE model with sex differences was selected for the BMI fluctuation in adolescence and BMI trajectory in early adulthood in both sexes.
Abbreviations: a<sup>2</sup>: proportion of total variance explained by additive genetic factors; e<sup>2</sup>: proportion of total variance explained by unique environmental factors; LL: lower limit; UL: upper limit

Table 3 displays the phenotypic, additive genetic and unique environmental correlations between the analysed pairs of traits. The BMI fluctuation in adolescence was only linked to BMI trajectories in early adulthood in females (r=0.18). Additionally, BMI in adolescence had significant phenotypic associations with BMI fluctuations in early adulthood in both males (r=0.16 – 0.30) and females (r=0.37 – 0.39). However, BMI trajectories in adolescence and BMI fluctuations in early adulthood only showed a significant phenotypic association in males (r=0.14). The association between BMI fluctuation in adolescence and early adulthood was only significant in males (r=0.26). Subsequently, we decomposed the significant correlations into genetic and environmental components. Genetic factors accounted for most of the phenotypic associations between BMI fluctuation and BMI trajectories: the additive genetic correlations (r_A_=0.14 for the associations between BMI fluctuation in adolescence and BMI trajectories in early adulthood; r_A_=0.16 – 0.51 for the associations between BMI in adolescence and early adulthood BMI fluctuation; r_A_=0.50 for the associations between BMI fluctuations in adolescence and early adulthood in males) were generally higher than the phenotypic correlations. However, in males, the association between BMI fluctuation in early adulthood and BMI trajectory in adolescence was entirely (r_E_=0.24) and the associations between BMI at wave 3 and early adulthood BMI fluctuation partly attributable to unique environmental factors (r_E_=0.22).

**Table 3.**
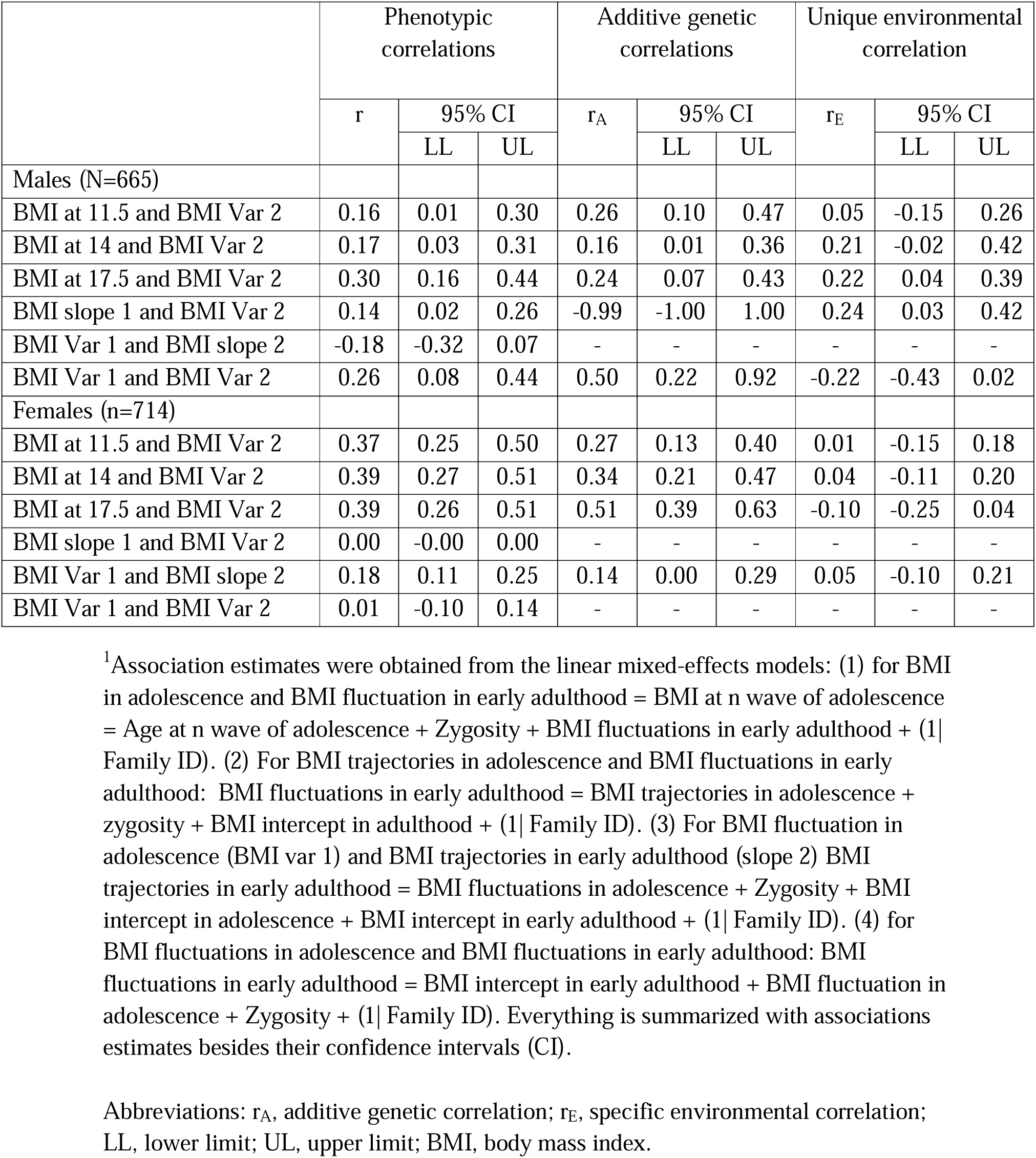
Phenotypic associations, additive genetic and unique environmental correlations of BMI, BMI trajectories and BMI fluctuation in adolescence (Var 1, slope 1) versus early adulthood (Var 2, slope 2) by sex, and specific ages.^1^.

|  | Phenotypic correlations |  |  | Additive genetic correlations |  |  | Unique environmental correlation |  |  |
| --- | --- | --- | --- | --- | --- | --- | --- | --- | --- |
|  | r | 95% CI |  | r<sub>A</sub> | 95% CI |  | r<sub>E</sub> | 95% CI |  |
|  |  | LL | UL |  | LL | UL |  | LL | UL |
| Males (N=665) |  |  |  |  |  |  |  |  |  |
| BMI at 11.5 and BMI Var 2 | 0.16 | 0.01 | 0.30 | 0.26 | 0.10 | 0.47 | 0.05 | -0.15 | 0.26 |
| BMI at 14 and BMI Var 2 | 0.17 | 0.03 | 0.31 | 0.16 | 0.01 | 0.36 | 0.21 | -0.02 | 0.42 |
| BMI at 17.5 and BMI Var 2 | 0.30 | 0.16 | 0.44 | 0.24 | 0.07 | 0.43 | 0.22 | 0.04 | 0.39 |
| BMI slope 1 and BMI Var 2 | 0.14 | 0.02 | 0.26 | -0.99 | -1.00 | 1.00 | 0.24 | 0.03 | 0.42 |
| BMI Var 1 and BMI slope 2 | -0.18 | -0.32 | 0.07 | - | - | - | - | - | - |
| BMI Var 1 and BMI Var 2 | 0.26 | 0.08 | 0.44 | 0.50 | 0.22 | 0.92 | -0.22 | -0.43 | 0.02 |
| Females (n=714) |  |  |  |  |  |  |  |  |  |
| BMI at 11.5 and BMI Var 2 | 0.37 | 0.25 | 0.50 | 0.27 | 0.13 | 0.40 | 0.01 | -0.15 | 0.18 |
| BMI at 14 and BMI Var 2 | 0.39 | 0.27 | 0.51 | 0.34 | 0.21 | 0.47 | 0.04 | -0.11 | 0.20 |
| BMI at 17.5 and BMI Var 2 | 0.39 | 0.26 | 0.51 | 0.51 | 0.39 | 0.63 | -0.10 | -0.25 | 0.04 |
| BMI slope 1 and BMI Var 2 | 0.00 | -0.00 | 0.00 | - | - | - | - | - | - |
| BMI Var 1 and BMI slope 2 | 0.18 | 0.11 | 0.25 | 0.14 | 0.00 | 0.29 | 0.05 | -0.10 | 0.21 |
| BMI Var 1 and BMI Var 2 | 0.01 | -0.10 | 0.14 | - | - | - | - | - | - |

<sup>1</sup>Association estimates were obtained from the linear mixed-effects models: (1) for BMI in adolescence and BMI fluctuation in early adulthood = BMI at n wave of adolescence = Age at n wave of adolescence + Zygosity + BMI fluctuations in early adulthood + (1| Family ID). (2) For BMI trajectories in adolescence and BMI fluctuations in early adulthood: BMI fluctuations in early adulthood = BMI trajectories in adolescence + zygosity + BMI intercept in adulthood + (1| Family ID). (3) For BMI fluctuation in adolescence (BMI var 1) and BMI trajectories in early adulthood (slope 2) BMI trajectories in early adulthood = BMI fluctuations in adolescence + Zygosity + BMI intercept in adolescence + BMI intercept in early adulthood + (1| Family ID). (4) for BMI fluctuations in adolescence and BMI fluctuations in early adulthood: BMI fluctuations in early adulthood = BMI intercept in early adulthood + BMI fluctuation in adolescence + Zygosity + (1| Family ID). Everything is summarized with associations estimates besides their confidence intervals (CI).
Abbreviations: r<sub>A</sub>, additive genetic correlation; r<sub>E</sub>, specific environmental correlation; LL, lower limit; UL, upper limit; BMI, body mass index.

Finally, we examined the association between PRS_BMI_ with BMI trajectories and BMI fluctuation in adolescence and early adulthood (Table 4). In males, PRS_BMI_ was associated with BMI fluctuation (β=0.13) and trajectory in early adulthood (β=0.11). In females, statistically significant associations were found with BMI fluctuation in adolescence (β=0.21) and early adulthood (β=0.23), as well as with BMI trajectory in early adulthood (β=0.16).

**Table 4.** Associations between polygenic risk score of BMI (PRS_BMI_) with body mass index (BMI) trajectories and fluctuation in adolescence and early adulthood by sex.^1^.

|  | Males (n=665) |  |  | Females (n=714) |  |  |
| --- | --- | --- | --- | --- | --- | --- |
| | $\beta$ | 95% CI | | $\beta$ | 95% CI | |
|  |  | LL | UL |  | LL | UL |
| BMI fluctuation in adolescence | 0.13 | -0.11 | 0.38 | 0.21 | 0.04 | 0.37 |
| BMI fluctuation in early adulthood | 0.18 | 0.06 | 0.32 | 0.23 | 0.05 | 0.38 |
| BMI trajectory in adolescence | 0.01 | -0.00 | 0.03 | 0.00 | -0.01 | 0.01 |
| BMI trajectory in early adulthood | 0.11 | 0.04 | 0.30 | 0.16 | 0.02 | 0.27 |

<sup>1</sup>Associations obtained from linear mixed-effect models (1) BMI trajectories in adolescence or early adulthood = $PRS_{BMI}$ + Zygosity + BMI intercept in concrete stage + Baseline age of concrete stage + (1| Family ID) for obtaining those between BMI trajectories in both adolescence and early adulthood and the polygenic risk scores and, (2) BMI fluctuation = $PRS_{BMI}$ + Zygosity + BMI intercept in concrete stage + (1| Family ID) for those between BMI fluctuation in adolescence and early adulthood and polygenic risk scores for BMI are summarised with association coefficients beside their confidence intervals (CI).
Abbreviations: LL, lower limit; UL, upper limit

## Discussion

The current study examines the associations between BMI, BMI trajectories and BMI fluctuations in adolescence with the BMI trajectories and fluctuation in early adulthood. Firstly, we found that BMI in adolescence is associated with BMI fluctuation in early adulthood. This indicates that greater BMI in adolescence is associated with greater BMI fluctuation (more erratic weight changes) in early adulthood. The associations in the early (11.5 years) and middle (14 years) stages of adolescence were fully attributable to genetic factors. In contrast, the association observed in the late stages of adolescence in males was influenced by both genetic and environmental factors.

Secondly, in males the BMI trajectories in adolescence are related to BMI fluctuation in adulthood, indicating that greater weight gain in adolescence is associated with more BMI fluctuation in adulthood. This association was found to be fully influenced by environmental factors. Moreover, the fluctuation in BMI in adolescence is associated with fluctuation in early adulthood in males, indicating that greater fluctuation in BMI in adolescence is associated with greater fluctuation in adulthood. This association was driven exclusively by genetic factors. Finally, the BMI fluctuation in adolescence was associated with BMI trajectories in early adulthood in females, indicating that the more BMI fluctuates in adolescence the more pronounced the weight gains in early adulthood are. This association was also explained purely by the genetic factors. Our results on the role of genetic factors behind BMI and its fluctuations were supported by the association between PRS_BMI_ with BMI trajectories and fluctuations in early adulthood. The results indicate that genetic factors influencing BMI variation may also contribute to the definition of BMI trajectories and fluctuation.

Previous studies have reported associations between BMI in adolescence and BMI in adulthood (31, 32), as well as between BMI trajectories in adolescence and BMI in adulthood (31, 33). Both genetic and environmental correlations explain these phenotypic associations (11, 34, 35). When interpreting these associations, it is noteworthy that during adolescence, trajectories in BMI reflect changes in both height and weight, whereas in adulthood the trajectories in BMI are purely explained by weight change, primarily the accumulation of fat mass. Moreover, childhood and prepubertal BMI, as well as other obesity markers, are associated with earlier onset puberty, which may impact these associations (36, 37). It is known that earlier pubertal timing predicts higher BMI and increased risk of obesity in adulthood (38).

Furthermore, the associations between prepubertal or childhood BMI with puberty timing, as well as between puberty timing and adulthood BMI and obesity, are explained by both genetic and environmental factors (39, 40). Thus, the genetic factors associated with puberty may contribute to the association between BMI fluctuation in adolescence and weight gain in adulthood.

The heritability estimates of BMI fluctuations in adolescence were found to be high in males and low in females. In early adulthood, females presented moderate heritability, while males presented a low estimate. These findings indicate that genetic factors influence BMI fluctuations in adolescence, but the role of genetic factors diminishes in early adulthood for males while the opposite occurs for females. However, genetic factors continue to exert an influence in early adulthood in both sexes. Regarding BMI trajectories, heritability estimates are high in both life stages and both sexes. High heritability estimates of BMI have been reported previously in adolescence (41, 42) and adulthood (43, 44) (0.80 – 0.85 for adolescence and 0.57 – 0.72 for adulthood).

Additionally, BMI trajectories in adulthood have previously been shown to be heritable in another Finnish twin cohort (13). However, to the best of our knowledge, our study is the first to report the heritability estimates for BMI fluctuations.

The current study has both strengths and weaknesses. The most notable strength is the use of longitudinal population-based data spanning from early puberty to early midlife over a period of more than two decades and five measurement points, along with information not only on twins’ status but also on the genetic susceptibility of high BMI (obtained from DNA sample) which allows us to use two methods to analyse the role of genetics, each based on different assumptions. However, the sample size decreased due to selecting only participants with BMI measurements at all ages, potentially reducing statistical power and increasing uncertainty in our estimates. The number of measurement points does not fully cover the BMI trajectories and therefore also the fluctuations. This may lead to an underestimation of the true variations, especially among those with a lot of weight cycling. This underestimation may be more pronounced in adulthood, considering the 13-year time gap between the last 2 measurements. However, to the best of our knowledge, there are limited observational cohorts that have measured BMI values over a sufficiently long period of time with short time periods between BMI measurements to more precisely capture weight cycling (e.g., Nurses’ Health Study (NHS) (45) and Health Professionals Follow-up Study (HPFS) (46)). Linear mixed-effect models used to estimate BMI trajectories do not capture non-linear patterns in BMI trajectories. However, capturing non-linear patterns with a limited number of BMI measurements per individual is challenging.

Another limitation that can be mentioned is the lack of control for potential confounders such as diet or physical activities among others.

In conclusion, the current study provides evidence of a shared genetic background between BMI, BMI trajectories, and BMI fluctuations in adolescence and early adulthood using both twin modelling and polygenic risk scores. These associations emphasize the significance of adolescence in understanding BMI development in early adulthood, as temporal changes and fluctuations in BMI in adolescence are related to these traits in adulthood. This knowledge is important for identifying individuals at a higher risk of weight gain in early adulthood. However, while these findings are promising, it would be advantageous to confirm them in other populations to improve the generalizability.

## Data Availability Statement

The FinnTwin12 (FT12) data is not publicly available due to the restrictions of informed consent. However, the FT12 data is available through the Institute for Molecular Medicine Finland (FIMM) Data Access Committee (DAC) for authorized researchers who have IRB/ethics approval and an institutionally approved study plan. To ensure the protection of privacy and compliance with national data protection legislation, a data use/transfer agreement is needed, the content and specific clauses of which will depend on the nature of the requested data.

## Code availability

All R scripts are available from the corresponding author.

## Author and contributions

AO created the study design, conducted the analyses, and drafted the manuscript. JK and SA collected the data. AJ, JG, GD, SA, JK and KS participated in the interpretation of the data and critically drafted the manuscript. All authors approved the submitted version of the manuscript.

