## Supplementary tables for "Genetic contributions to BMI fluctuation and its associations with BMI and its trajectories over adolescence and early adulthood: a 25-year follow-up longitudinal study of Finnish twins"

**Supplementary table 1:** Correlations between the body mass index fluctuation in adolescence and adulthood obtained through delta model (1) and linear mixed effects model (2).

|  | Males (N=665) | | | Females (N=714) | | |
| --- | --- | --- | --- | --- | --- | --- |
|  | r | 95% CI | | r | 95% CI | |
|  |  | LL | UL |  | LL | UL |
| BMI fluctuation in adolescence (1)- BMI fluctuation in adolescence (2) | 0.79 | 0.74 | 0.83 | 0.78 | 0.74 | 0.82 |
| BMI fluctuation in adulthood (1)- BMI fluctuation in adulthood (2) | 0.91 | 0.88 | 0.93 | 0.95 | 0.93 | 0.97 |

Abbreviations: BMI: Body mass index; LB: Lower bound; UB: Upper bound; CI: Confidence interval.

Methodological note: To study how the method used in the current study for obtaining the variability of BMI fits with real variability, delta slope method was carried out between two consecutives waves (wave 1-2 and wave 2-3 for adolescence while wave 3-4 and wave 4-5 for adulthood) obtaining a delta slope estimate for each individual in each of the analysis and then obtaining the variance of these estimates (Variance of delta slopes obtained from wave 1-2 and 2-3 in adolescence; Variance of delta slopes obtained from wave 3-4 and 4-5 in adulthood).

Results of Pearson correlation analysis carried out between the two ways of obtaining the BMI fluctuation, (1) by applying the formula *y = xn + m*, where *n* is the BMI trajectory measured in kg/m^2^ per year (i.e., slope), *m* is the baseline BMI measured in kg/m^2^ (i.e., wave 1 for adolescence and wave 3 for early adulthood), and *x* is the difference between the age of interest and the baseline age (wave 1 for adolescence and wave 3 for early adulthood). After that squared differences between observed and expected BMI were calculated at each age. Finally, the total BMI fluctuation in adolescence and early adulthood was obtained for each individual by calculating the average of the squared difference values (using the obtained values at waves 1, 2 & 3 for adolescence and the obtained values at waves 3, 4,& 5 for early adulthood) while (2) is based on obtaining delta slopes between each wave (wave 1-2, wave 2-3, wave 3-4 and wave 4-5), and obtain the mean value of slope (mean value of slopes obtained in wave 1-2 and 2-3 for adolescence; mean value of slopes obtained in wave 3-4 and 4-5) for adulthood. Finally, the formula ((x_1_− x̅)^2^+(x_2_− x̅)^2^)/n (x_1_ and x_2_ are the two slopes estimates; x̅ is the mean value of x_1_ and x_2_; 2 is the number of slopes used in each phase, in this case 2) was applied to obtain the variance in each stage (adolescence and adulthood).

**Supplementary table 2:** Intraclass correlations for the body mass index variability and trajectories in both adolescence (from ages 11.5 to 17.5) and adulthood (from ages 24 to 37 years) by sex and zygosity.^1^

|  | Males | | | | | | Females | | | | | | Opposite-sex DZ twins | | |
| --- | --- | --- | --- | --- | --- | --- | --- | --- | --- | --- | --- | --- | --- | --- | --- |
|  | MZ | | | DZ | | | MZ | | | DZ | | |  | | |
|  | r | 95% CI | | r | 95% CI | | r | 95% CI | | r | 95% CI | | r | 95% CI | |
|  |  | LL | UL |  | LL | UL |  | LL | UL |  | LL | UL |  | LL | UL |
| BMI var 1 | 0.60 | 0.52 | 0.67 | 0.12 | 0.00 | 0.23 | 0.43 | 0.33 | 0.51 | 0.71 | 0.65 | 0.76 | 0.30 | 0.22 | 0.37 |
| BMI var 2 | 0.34 | 0.10 | 0.54 | 0.42 | 0.11 | 0.66 | 0.60 | 0.45 | 0.72 | 0.56 | 0.36 | 0.71 | 0.01 | -0.18 | 0.21 |
| Slope 1 | 0.62 | 0.54 | 0.69 | 0.28 | 0.17 | 0.38 | 0.57 | 0.49 | 0.64 | 0.27 | 0.16 | 0.38 | -0.01 | -0.09 | 0.07 |
| Slope 2 | 0.59 | 0.40 | 0.73 | 0.55 | 0.27 | 0.75 | 0.63 | 0.48 | 0.74 | 0.38 | 0.15 | 0.58 | 0.20 | -0.00 | 0.38 |

^1^Intraclass correlations (mixed-effects ANOVA, decomposing variability into between- and within-subjects components) for BMI variability in adolescence (from ages 11.5 to 17.5 years) (i.e. BMI var 1) and adulthood (from ages 24 to 37 years) (i.e. BMI var 2), and the trajectories in BMI in adolescence (from ages 11.5 to 17.5 years) (i.e. Slope 1) and adulthood (from ages 24 to 37 years) (i.e. Slope 2) are summarized by sex and zygosity with correlation coefficients besides their 95% confidence interval. All the correlations are highly significant (all p<0.005).

Abbreviations**:** MZ: monozygotic; DZ: dizygotic; LL: lower limit; UL: upper limit.

**Supplementary table 3:** Model fit statistics of BMI variability and trajectories and underlying factors comparing different genetic models.^1^

|  | Full ACE model (reference model) | | ACE model without sex-specific genetic effect^2^ | | ACE model with same parameter estimates for boys and girls^3^ | | Full AE model^4^ | | AE model with same parameter estimates for boys and girls^5^ | | AE model without sex-specific genetic effect^6^ | |
| --- | --- | --- | --- | --- | --- | --- | --- | --- | --- | --- | --- | --- |
|  | 2LL | d.f | Δ -2 LL | p value | Δ -2 LL | p value | Δ -2 LL | p value | Δ -2 LL | p value | Δ -2 LL | p value |
| BMI var1 | 16542 | 3648 | 3.91 | 0.04 | 46.22 | <0.0001 | 56.50 | <0.0001 | 6.50 | 0.03 | 0.11 | 0.73 |
| BMI var2 | 7500 | 1319 | 0 | 0.97 | 20.87 | <0.0001 | 11.16 | <0.0001 | 17.19 | <0.0001 | 4.71 | 0.03 |
| Slope 1 | -3756 | 3648 | 13.93 | <0.0001 | 43.34 | <0.0001 | 0.13 | 0.93 | 43.21 | <0.0001 | 116.48 | <0.0001 |
| Slope 2 | -2242 | 1319 | 0 | 0.98 | 18.18 | <0.0001 | 5.34 | 0.05 | 15.11 | <0.0001 | 35.43 | <0.0001 |

^1^Full ACE model is compared against the rest of the models displaying the -2 log likelihood and degrees of freedom for the reference model and the differences in -2log likelihood and the p value in the models compared with the reference one. ^2^Compared to full ACE model (Δ d.f. 1); ^3^Compared to ACE model without sex-specific genetic effect (Δ d.f. 2); ^4^Compared to full ACE model (Δ d.f. 2); ^5^Compared to the full AE model (Δ d.f. 1); ^6^Compared to the AE model without sex-specific genetic effect (Δ d.f. 1).

Abbreviations: -2LL (-2 log-likelihood); d.f. (degrees of freedom); Δ (change); ACE (additive genetic/ shared environment/ unique environment) model; AE (additive genetic/ unique environment) model.
